# Mobility and non-household environments: understanding dengue transmission patterns in urban contexts

**DOI:** 10.1101/2024.05.28.24308061

**Authors:** Víctor Hugo Peña-García, Bryson A. Ndenga, Francis M. Mutuku, Donal Bisanzio, A. Desiree LaBeaud, Erin A. Mordecai

## Abstract

Households (HH) have been traditionally described as the main environments where people are at risk of dengue and other arbovirus infections. Mounting entomological evidence suggests a larger role for environments other than HH. Recently, an agent-based model (ABM) estimated that over half of infections occur in non-household (NH) environments such as workplaces, markets, and recreational sites. Despite the inferred importance of NH sites, we do not yet know how their urban spatial configurations, and human and vector mobility between them, affects their role in dengue transmission. To address this gap, we expanded an ABM calibrated with field data from Kenya to examine movement of people and vectors under different spatial configurations of buildings. We assessed the number of people traveling between HH and NH and the distances traveled, in three urban configurations: NH distributed randomly (scattered), concentrated in a single center, or clustered in multiple centers. Across simulations, the number of people moving was the most influential variable, with higher movement between HH and NH increasing case numbers. The number of cases was also higher when NH were scattered compared to centered or clustered. Intriguingly, the distance people traveled from HH to NH had little effect on dengue burden but influenced the spatial clustering of infections. These findings underscore the role of NH as major spreaders of infections between HH and NH environments, and the importance of human movement in driving dengue dynamics.

**Author summary:** Recent evidence describes a major role of non-household (NH) environments in dengue transmission. This new knowledge implies that spatial distribution of these locations and the movement of humans among them has implications in dynamics of transmission. However, these haven’t been evaluated neither together nor considering a differential role in transmission of urban environments. We modified a previously informed agent-based model to include spatial variables by assessing three different urban conformations, i.e. when NH are randomly distributed (scattered), centered, or grouped in differed clusters (clustered). On these, we also evaluated the movement of people from households (HH) to NH (both distance and number of individuals). Our findings, which includes a higher burden of dengue when NH are scattered and at higher levels of human movement, are aligned with the idea that infections are mainly happening in NH mediated by human mobilization. Infected people reach the households where local subpopulation of vector spread the virus to remaining inhabitants. This work highlights the role of NH and human mobility in dengue transmission.

## Introduction

Dengue is a vector-borne disease prevalent and on the rise in most of the tropical and subtropical regions around the globe, with introduced and locally acquired cases being reported in non-endemic areas like USA and Europe [1, 2]. The main vector, *Aedes aegypti,* is highly anthropophilic, found very close to human environments, and impacting the public health of urban environments [3, 4].

Historically, the main strategy to control the disease has been to reduce vector-human contact by reducing the size of mosquito populations. Conventional wisdom is that for effective control, these activities should be focused on households (HH) as the main environment where transmission is happening [5, 6]. However, some studies have suggested that locations other than households might have an important role because of a significant presence of mosquitoes [7–13] and infected vectors [14, 15] and the fact that this vector bites during the day [16–19], when people may be outside the home. In a recent study, we used an agent-based model (ABM) to quantify the number of infections in different types of urban spaces based on fieldwork mosquito collections and seroincidence data and estimated that over half of infections are happening in non-household (NH) environments, where the main high-risk spaces are workplaces and markets/shops [20].

These results have implications for dengue epidemiology since the high flux of people through NH suggests that these spaces can contribute to the spread of infections. In this way, the total number of infections can be affected by the distribution in space of NH and the movement of people between HH and NH. However, a key knowledge gap is how these intra-urban dynamics vary with spatial configuration and mobility levels [21–24], though Massaro and colleagues used mobile phone data to get estimates for movement between workplaces [25].

Building on a previous result showing the importance of NH for dengue transmission, we use an ABM to address this knowledge gap. In particular, what role does spatial configuration of NH spaces play, along with the extent to which people and mosquitoes move between spaces, in determining dengue dynamics? To address this, we modified a previously published ABM [20] to make it spatially explicit by assigning coordinates that mimic different urban conformations and evaluated different scenarios of movement of people and vectors. We then assessed how these variables affect the burden of dengue and the spatial patterns to understand urban-level transmission dynamics.

## Methods

### Model overview

To achieve the aims of this study, we modified the ABM previously used to describe the importance of HH and NH in transmission [20]. The model was developed to quantify the relative contribution of five different types of NH (workplaces, markets or shops, recreational, religious, and schools) and HH to dengue burden. The model development and calibration were based on data from two Kenyan cities: Kisumu in the west and Ukunda on the coast [20]. Here, we focus on parameters calibrated to Kisumu, although additional results including dynamics from Ukunda are found in supplementary material.

The model represents the movement of people between HH and two different types of NH locations. The first are daily-commuting locations where individuals attend daily and meet with the same individuals, including schools (average number of students per school set to be 360 [26]) and workplaces (average number of workers per workplace is 19 [27]). The assignment of each individual as student, worker, or neither depends on their age and the age-specific proportion of individuals in such role according to governmental information [26–28]. The second type of NH locations are randomly assigned locations for which both the number and identity of people who visit them is randomly defined every day, including markets, shops, recreational, and religious spaces. Movement between HHs (categorized as “visit”) is also included in the model with a daily probability for incoming visits of 0.1, according to information extracted from a human movement survey conducted in Kenya [20].

Epidemiologically relevant locations where individuals move are those with presence of mosquitoes. Thus, based on vector surveys conducted over two years of fieldwork previously published [29], NH and HH environments were assigned to have mosquito presence or absence based on observed prevalence of mosquitoes. Population dynamics of vectors were modeled at the building level, whereby sub-population dynamics are driven by site-specific features—particularly the availability and capacity of water containers as well as numbers of individuals present—as informed by field surveillance data. These dynamics as well as infection dynamics of vectors are also determined by temperature by using functions previously described and widely used elsewhere [30–32].

Initial baseline prevalence of dengue was set to 0.08%, estimated from previous studies reporting age-structured seroprevalence [33] with an incidence rate per year estimated 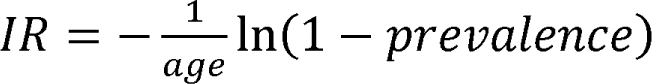. Transmission events happen in those locations where infected vectors bite susceptible humans or vice versa. Mosquitoes bite depending on both temperature-dependent biting rate and the probability of having a successful vector-human encounter, which depends on the amount of time that humans spend in the location. This information was obtained from a human movement survey where the time and frequency that individuals spend in different types of locations were recorded [20]. Infection status of mosquitoes can be either susceptible, exposed, or infected while humans can be either susceptible, exposed, infected, or recovered and (temporarily) immune. The time that mosquitoes spend as exposed depends on temperature (extrinsic incubation period) and is determined in the model by equations reported previously by Mordecai and colleagues [30]. Once infected, mosquitoes remain in this stage until death, which is evaluated daily following a temperature-dependent death rate (probabilistic implementation within the model is described in supplementary material). Humans remain susceptible until they are bitten by infected mosquitoes and moved to a latent stage where they remain for five days. Then, the individual is moved to the infectious stage, which lasts seven days, before moving to the recovered stage. Since the model does not explicitly represent dynamics of different serotypes, waning immune protection was based on Sabin’s classic studies describing the loss of complete heterotypic protection after roughly three months [34, 35] to set a return to susceptible after 100 days in recovered status. The number of infections in each location—number of infectious bites from mosquito to human leading to successful infection within a given physical location— is recorded daily. Statistics about the total number of infections and locations are reported weekly. The model simulates transmission dynamics happening for 731 days (comprising temperature conditions between January 1^st^ of 2020 until December 31^st^ of 2021) and results are shown as a distribution of the number of infections over 200 simulations. The model and modifications described in this work were coded in Julia language (v1.10.0) and simulations were run on Sherlock computing cluster (Stanford Research Computing Center). A more complete and detailed description of the model can be found at [20].

### Spatial variables

The original model was not spatially explicit and hence the movement of individuals was assumed to be totally random, with equal probability of visitation for all NH locations within a given NH type and by any given individual. As such, the resulting infections arise from complete mixing of individuals among structures, which is not realistic and does not capture local, intra-urban spatial phenomena. To include mobility-associated variables and describe such local phenomena, we made the model spatially explicit, where the transmission dynamics previously described are still happening but in a non-random, distance-dependent manner. To achieve this, we constructed an artificial coordinate system and tested different urban configurations using the same spatial framework. In this way, we can assure that differences are due to the building designation as HH or NH and not by spatial disparities coming from different sets of spatial coordinates. The model considers a synthetic population of 20,160 people, so the total municipality areas were rescaled to fit the total number of structures of the virtual populations while considering similar densities. Spatial coordinates were randomly generated to create a synthetic settlement and assigned to each structure of the population. With the aim of capturing different intra-urban conformations, coordinate assignments were done based on either “scattered” (randomly distributed), “centered” (majority of NH concentrated in the center of the city in a single cluster), or “clustered” (majority of NH concentrated in three clusters) configurations (Fig 1).

**Fig 1:**
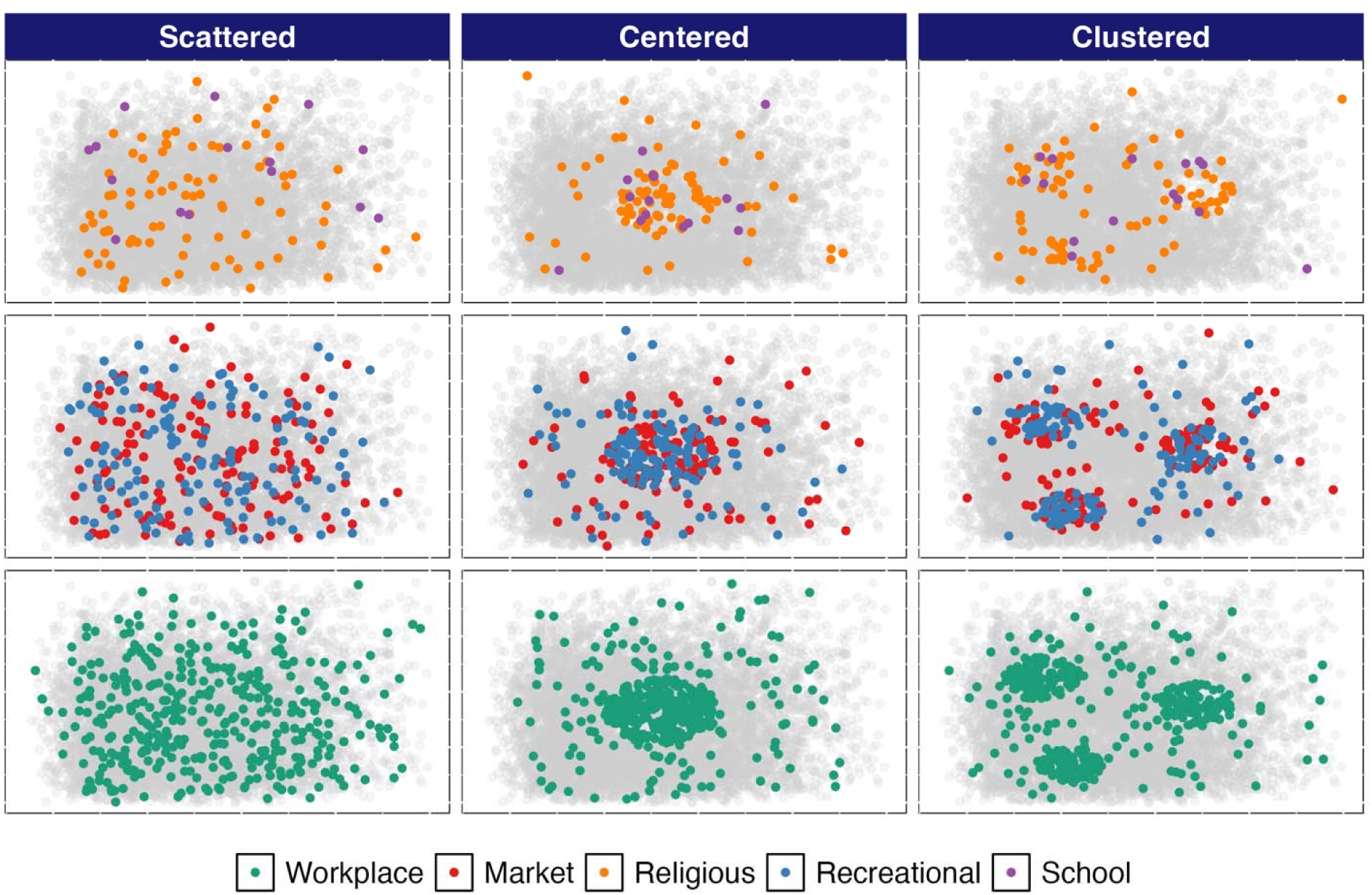
**Spatial distribution of NH for three different urban conformations to be tested:** scattered (NH randomly distributed in space), centered (majority of NH are clustered in the center), and clustered (majority of NH are grouped in three clusters). NH are shown with colors while HH are displayed as light gray points. Religious (orange) and schools (purple) are represented in the first row; markets/shops (red) and recreational spaces (blue) in middle row; and the most abundant NH, workplaces (green), are represented in the lower row.

### Movement of people

We included two movement-related variables: the distance from each household (HH) to the nearest non-household (NH) locations and the number of people moving. To control movement distances, we applied three treatments: (1) limiting attendance to the nearest NH locations (assigned as distance zero), (2) allowing attendance to NH locations at least 500 meters away, and (3) allowing attendance to NH locations at least 1000 meters away from each HH. These treatments were applied across the three different urban configurations. The results are entirely based on computational simulations; no experiments involving humans were conducted.

In the clustered or centered configurations, assigning the closest NH locations to HHs would primarily select NH sites at the periphery of each cluster, potentially leading to biased representations. To address this and make it equivalent to scattered configuration, we generated a list of NH locations sorted by proximity for each HH and allocated the closest NHs based on the number of inhabitants in the HH. For example, if a HH had four inhabitants, the four closest NHs were assigned to it. This approach ensured a more representative distribution across all HHs.

The number of people visiting NH was simulated at three levels. First, we included the same levels previously described in the model, categorized as 100% mobility [20]. This treatment includes all students and workers attending their respective school and workplace, and random-attendance locations (religious, markets/shops, and recreational). Unfortunately, no data were available on the number of individuals visiting these types of locations. We anticipated significant variability due to the influence of several unmodeled factors. To account for this uncertainty, we relied on discussions with local residents to estimate a range of possible visitor numbers. We then applied a uniform distribution, with a minimum of 10 and a maximum of 70 individuals, to cover a broad spectrum of potential scenarios. The number of people visiting a given location is drawn from this uniform distribution on the interval [10,70] daily. We also evaluated a medium-mobility scenario by decreasing the number of people moving to school and workplaces to 50% and remaining NH locations to a uniform distribution with parameters minimum = 5 and maximum = 35. Finally, we include low-mobility scenario of 20% for school and workplaces and a uniform distribution with parameters minimum = 2 and maximum = 14 for random-attendance locations.

### Movement of mosquitoes

In line with the inclusion of spatial variables and human movement, we included the movement of vectors. There were no data available to characterize the rate of movement of mosquitoes into new areas. As a result, we parameterized such movement by considering two variables—availability of both breeding and blood-feeding resources—to estimate a baseline migration probability for the number of mosquitoes moving from given locations.

The model includes a local density-dependent function that allows for the mosquito population to grow in a location-specific way, representing dynamics previously described for a fragmented environment [36]. The function, as previous described [20], depends on temperature (peaking at 29°C) and the density of immatures according to water availability, as follows:

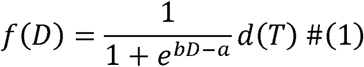

Where

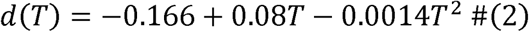

In this equation *a* and *b* are calibrated parameters, *T* is environmental temperature in degrees Celsius, *D* is the larval density expressed as the ratio of the number of larvae to liters of water available for breeding in the structure, and *d*(*T*) is the term describing the temperature-dependence of population growth.

By using this function, the growth of the mosquito sub-population (in a given building) depends on the amount of water resources available in the structure. In this way, when the mosquito population has grown so the water resources are depleted (reaching the carrying capacity), the mortality of immatures is higher and the probability for a given mosquito to migrate out of the building increases (details provided in supplementary material).

Additionally, we considered for migration the possibility that human blood availability is not sufficient to support the mosquito subpopulation in a given building. For this purpose, the number of females that fed in each day was estimated by considering the number of females biting (*N_b_*) and the probability for those to have a successful feeding encounter with a human the same day (*P*(*bit*)). The latter is determined considering the number of individuals, the time they spend at the building, and the time a visitor spends in each location (details are provided in supplementary material). The final number of fed females (*N_F_*) is estimated by

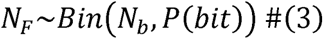

Thus, when the difference between the number of mosquitoes trying to bite and the actual number of mosquitoes that are fed is large, the probability of migration increases. Once a mosquito migrates, a new location is assigned by considering a dispersal kernel [37]. Following previous work [38], we used a lognormal function with the form

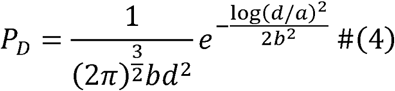

Where *d* is distance and both *a* and *b* are parameters to be estimated. We fitted a function by assuming a mean dispersal distance of 105.69 meters, as estimated for *Aedes aegypti* by [39].

### Spatial autocorrelation

By taking advantage of spatial features introduced in the model, we performed a spatial autocorrelation analysis to evaluate the level of clustering of dengue cases recorded in each simulation as a function of urban configuration and human and mosquito mobility. To do this, the location of each mosquito-human infectious encounter irrespective of the type—HH or NH—is recorded so a Global Moran’s *I* index [40] could be estimated at the end of the simulated period. Global Moran’s *I* ranges from -1 to +1, where -1 means totally dispersed location of cases while 1 represents a spatial distribution that is totally clustered (total separation between locations with dengue cases and those without cases). In this sense, the null hypothesis of this analysis is that dengue cases are randomly distributed in municipalities, represented by Moran’s *I* value of 0 [41]. The analysis was done for every simulation by using 1,000 permutations for inference in each of them. Analyses were done using the package SpatialDependence.jl implemented in Julia language (v 1.10.0). Code is available in the GitHub repository (https://github.com/vhpenagarcia/ABM_dengue) and have been archived within the Zenodo repository (https://doi.org/10.5281/zenodo.14036270).

## Results

### Burden of dengue is strongly affected by number of people visiting NH

We quantified the total number of cases after two years of transmission. When we simulated the epidemic under different human movement regimes, it was evident that the number of cases nonlinearly decreased as the number of people moving from HH decreased. For 100% human movement, irrespective of the urban conformation, we estimated a median of 4,228 cases (IQR: 3,025 – 4,921), which decreased to a median of 764 (IQR: 349 – 1,626) and 154 (IQR: 108 – 232) cases for 50% and 20% human movement, respectively (Fig 2 and Table S1) (all results are derived from the model calibrated for Kisumu, Kenya; see supplementary material for further results for Ukunda, Kenya).

**Fig 2:**
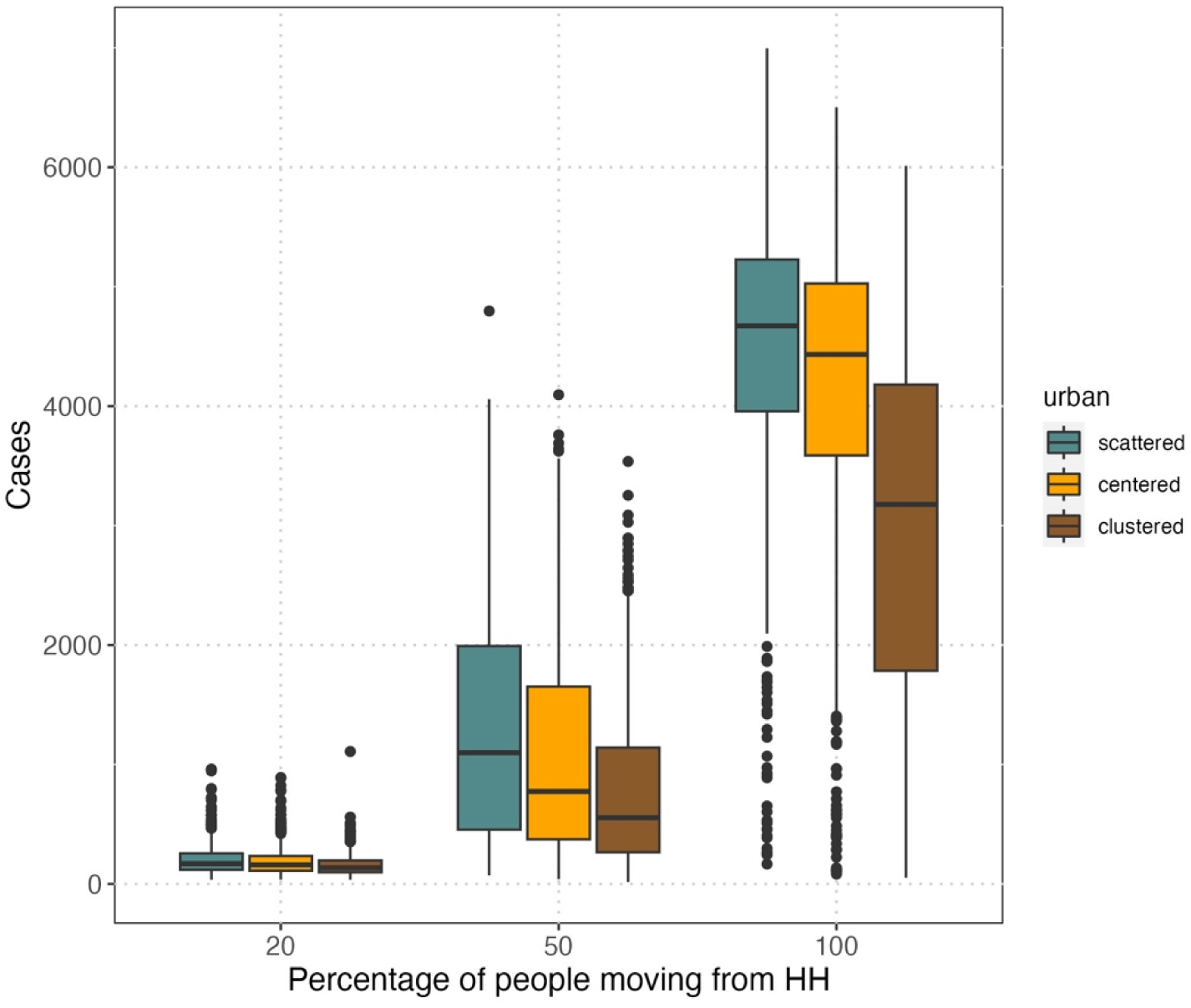
Increasing the number of people moving from HH to NH significantly increased the burden of dengue under three NH spatial distribution scenarios. Three levels of human movement were assessed (20%, 50%, and 100%) on three urban conformations (scattered, centered, or clustered). Boxplots show the distribution of the total number of infections for 200 runs of two-year simulations where median is the horizontal line, the filled box is the interquartile (IQR) range, the whiskers show the values above and under the IQR and no more than 1.5·IQR, and dots are representing values beyond this range.

Additionally, the scenario where NH locations are spatially randomly distributed produced more cases, though at all movement levels the interquartile ranges for different spatial configurations overlapped (Fig 2). At 100% movement, scattered conformation yielded a median of 4,672 (IQR: 3,956 – 5,227) while the centered and clustered scenarios produced, respectively, medians of 4,432 (IQR: 3,587 – 5,027) and 3,178 (IQR: 1,785 – 4,179) (Table S2). Trends were similar for the 50% and 20% mobility levels (Fig 2).

The relative role of NH versus HH environments increased with the overall level of mobility and hence transmission (Fig 3). At lower levels of movement (20%), the number of infections was slightly higher in HH than NH. However, at higher levels of movement (50-100%), the number of infections in NH was higher than HH. At 100% movement, irrespective of urban conformation, NH produced 67% of the cases, but this proportion decreased to 58.8% and to 42.3% at 50% and 20% of human movement, respectively (Fig 3 and Table S1). Mosquito movement had the opposite influence on transmission. Specifically, transmission decreased at higher levels of mosquito movement due to migration-mediated mortality (see supplementary results and discussion).

**Fig 3:**
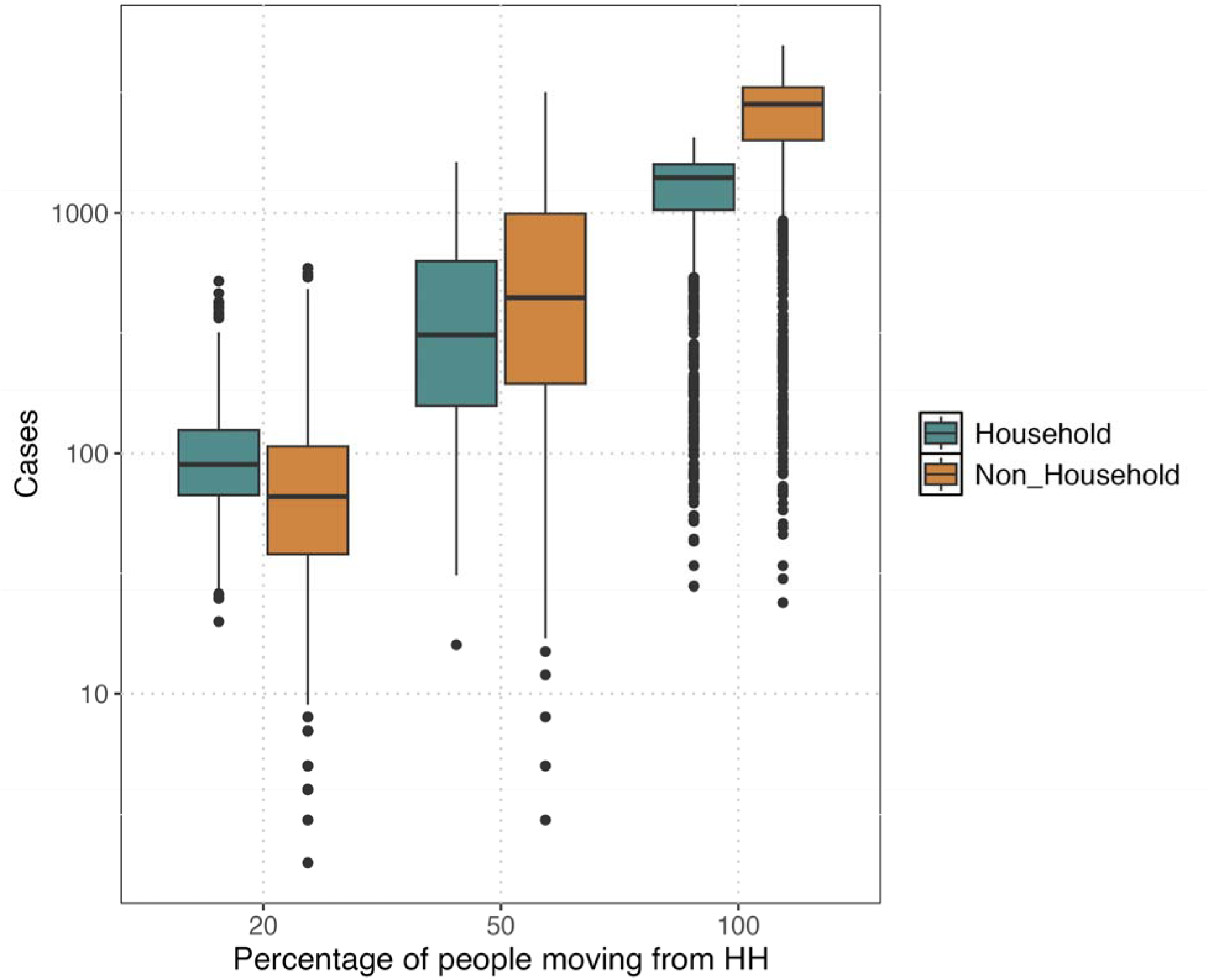
Number of infections are higher in NH than HH at high levels of human movement, but the HH contribution exceeds NH at low mobility and transmission levels. Three levels of human movement were assessed (20%, 50%, and 100%) on three urban conformations (scattered, centered, or clustered). Boxplots show the distribution of total number of infections for 200 runs of two-year simulations, spanning all three urban configurations, where median is the horizontal line, the filled box is the interquartile (IQR) range, the whiskers show the values above and under the IQR and no more than 1.5·IQR, and dots represent values beyond this range. The y-axis is represented in Log_10_ scale.

### Distance from HH to NH makes little difference in dengue burden but defines level of spatial structure

Varying the distance between HH and NH had only slight impacts on the total number of infections, with a slight increase in the number of cases with distance when NH are clustered (Fig 4 and Table S3). Besides these slight changes, differences among urban conformations were still evident (Fig 4).

**Fig 4:**
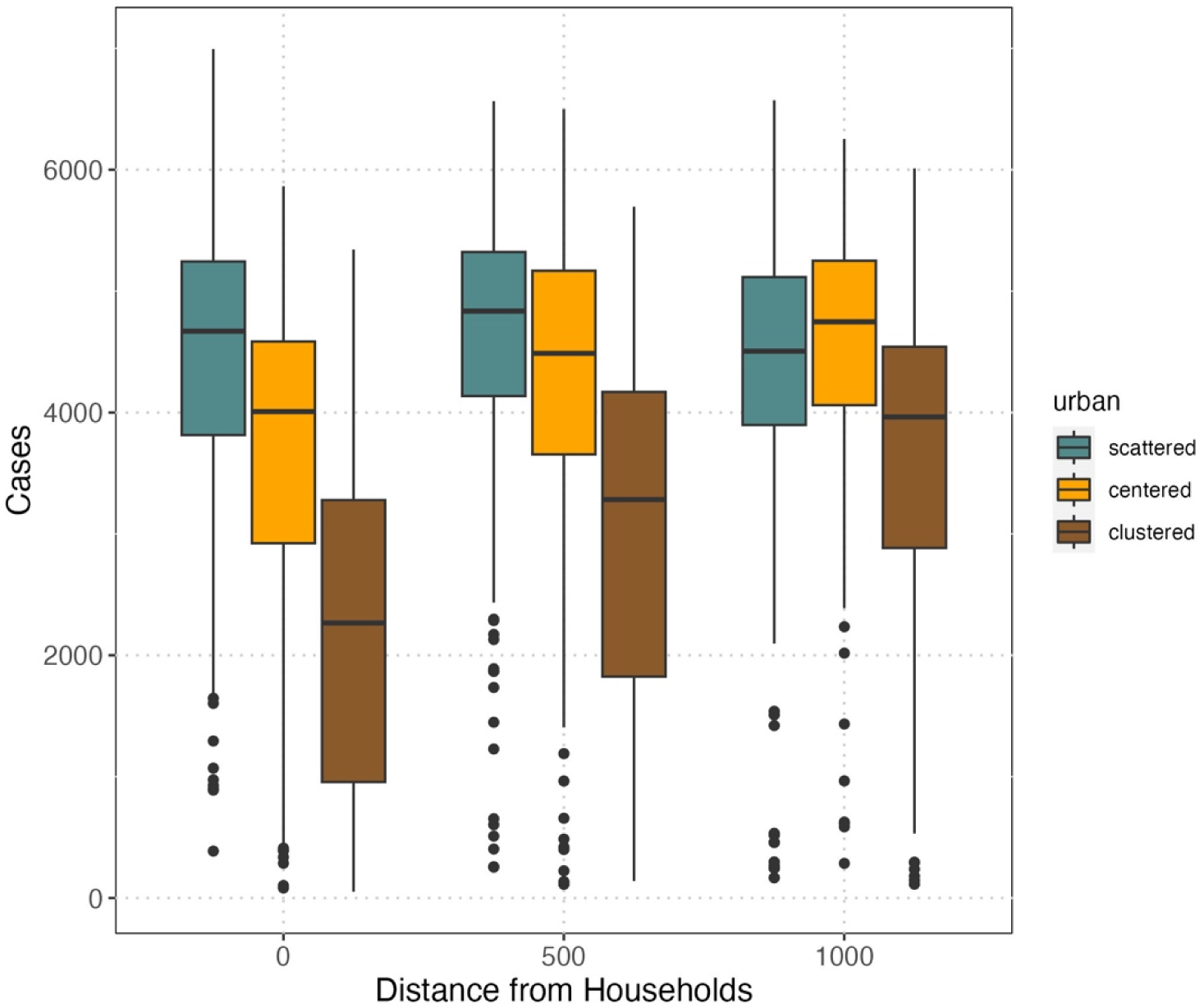
Different distance regimes people travel from HH to NH results in slight differences in cases and larger differences among configurations. Distance 0 means people only visit the closest NH location from HH. On the three urban configuration scenarios, three levels of distance from HH to NH were assessed (the closest [categorized as 0], at least 500 meters, and at least 1000 meters). Boxplots are showing the total number of infections after two-year simulation for 200 runs.

Intrigued by the apparent lower importance of human movement distance in transmission, we wanted to explore further by assessing the spatial structure of cases. Given that the number of people moving affected the number of cases, we evaluated the spatial structuring when distance traveled is considered as well. In general, we found that irrespective of urban conformation, when people moved short distances the level of spatial structure was higher, as expected (Fig 5). Similarly, the spatial structuring levels was also modified by the number of people moving, where lower levels of movement decreased Moran’s *I*, thereby making cases more dispersed (Fig S6).

**Fig 5:**
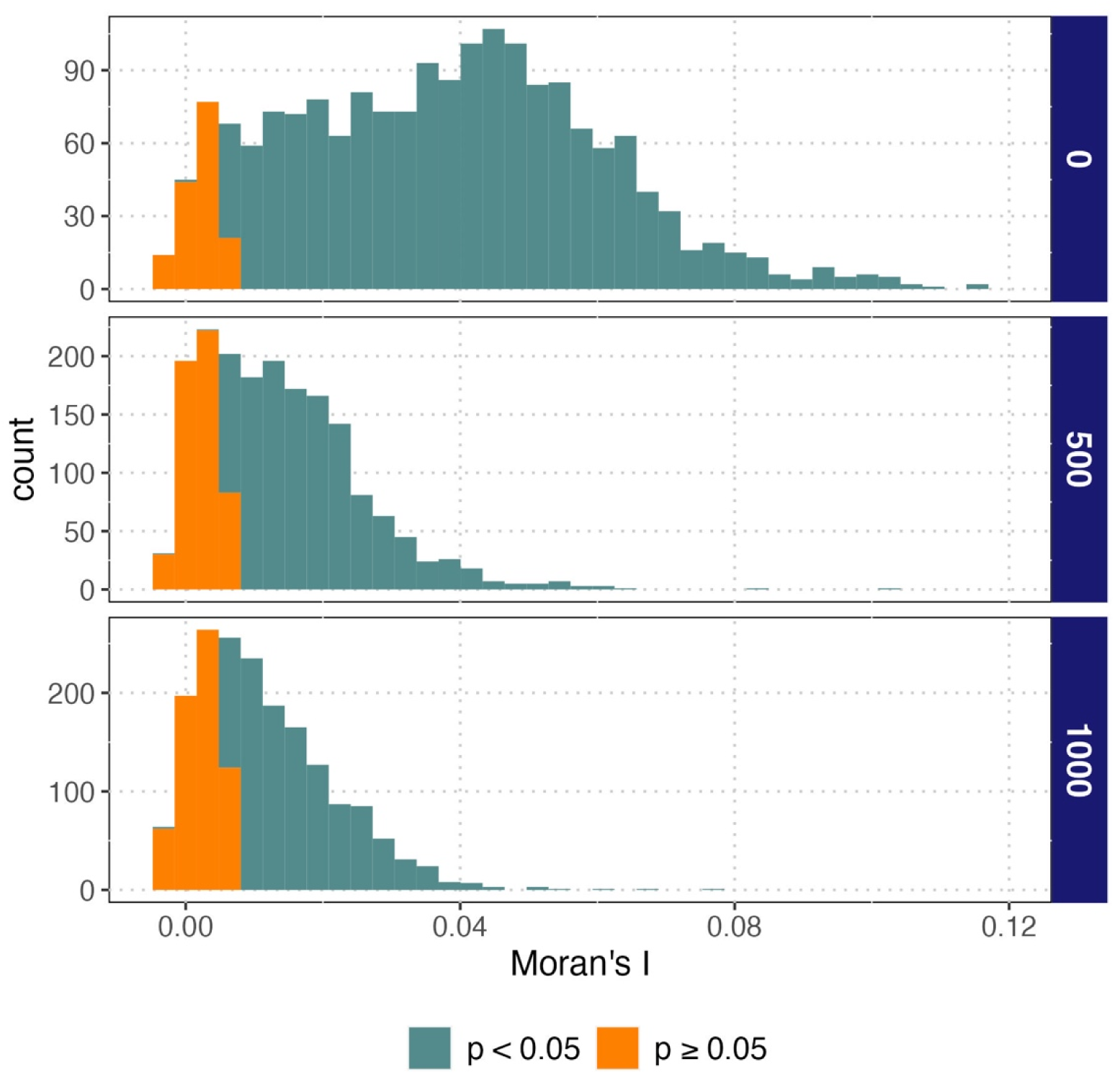
Spatial structuring decreased with increased distance of movement. Distribution of Moran’s I values for 200 simulations for each of the three distance regimes is shown. Values that are both significantly (α = 0.05; green) and non-significantly (orange) different from zero are displayed by color. Three levels of distance from HH to NH were assessed (the closest [categorized as 0], at least 500 meters, and at least 1000 meters) on three urban conformations (scattered, centered, or clustered).

## Discussion

Building on recent model results showing an important role of NH environments for dengue transmission and control, where over half of infections occur in NH environments, here we addressed how non-random mixing and mobility of humans and vectors affected dengue dynamics [20]. By extending the ABM to consider spatially-explicit urban conformations and movement levels, we showed that human movement is a primary driver of dengue dynamics, irrespective of urban spatial configuration. Further, we found qualitatively similar outcomes to those modeled for Kisumu for the coastal Kenyan city of Ukunda, presented in the supplementary material.

Among urban conformations, when NH spaces are scattered throughout the city it allows for closer connections to HH and therefore increased transmission. In this way, NH spaces serve as spreaders of infection since they are highly visited locations, which increases the chances of having a successful feeding encounter between humans and infected vectors. Once an individual is infected, the chances of infecting mosquitoes inside the household and in turn having another household inhabitant infected increases, generating local household chains of transmission.

For this reason, a lower number of individuals visiting NH locations nonlinearly reduces the burden of dengue; for example, a 50% reduction in mobility from 100% to 50% reduced cases by 82% (Fig 2). In this sense, when the number of people visiting NH decreases, the number of infections happening in these spaces also decreases until becoming roughly even with the number of infections in HH (Fig 3). These results are supported by previous reports showing that when COVID-19 pandemic lockdowns forced people to stay at home most of the time, the number of dengue cases was significantly reduced [24, 42, 43]. The idea of human movement affecting dengue transmission is not new and has been explored previously, especially by Stoddard and colleagues (2009), who focused their study on the movement of individuals between houses only [21]. However, here we highlight how human mobility interacts with NH spaces as drivers of transmission, which then disseminates within households. As a result, when control is focused on households, it prevents the spread of disease to the remaining household inhabitants but leaves broader foci of transmission active as long as NH transmission is not under control.

Distance from HH to NH was not as important as overall levels of human movement for dengue burden (Fig 4), suggesting that it is not how far people are traveling but the destination and total amount of movement. This is in line with previous work where a large, longitudinal study in Iquitos, Peru showed that human infection risk was mainly driven by individuals visiting locations with presence of infected vectors, irrespective of the distance [22]. It is important to note that our model does not account for movement times, which increase with distance (but note that even our largest range of movement, >1000m, is still very localized within a city’s limits, which allowed us to identify patterns but might be a limitation of such analysis). These NH spaces have been previously described to have mosquitoes [44] and hence represent some degree of risk for transmission when people are nearby. Unfortunately, we do not have data about the time people spend in NH locations. Equally, the limited information collected about the number of individuals within specific types of NH on a daily basis is a big limitation of the study. Although our simulations relied on information from observations and conversation with locals in Kisumu, Kenya, no quantitative activity space data exists in this setting. Although the results should not be dramatically different, it is likely that true patterns might be slightly different to what is reported in this work.

The distance that people travel to NH does, however, affect the urban spatial dynamics of transmission. By increasing people’s travel distances we are also increasing the mixing of individuals. Distance traveled affects the level of clustering of cases, which is a measure of the level of spatial dependence of cases and hence of how cases are unevenly distributed in space (Fig 5) [41]. The role of mobility between HH and NH in driving the clustering of cases is something that has not been explored before and deserves further exploration to understand its implication for disease control program design.

Although urban spatial configuration had subtle effects on the number and spatial structure of dengue infections, human movement between HH and NH had a much larger impact, with an 82% decline in cases as the number of people moving decreased from 100% to 50%. Together, these results reflect the importance of NH and human mobility between NH and HH spaces in dengue epidemiology. This underscores the importance of vector control in NH spaces, which is not currently implemented in many dengue endemic regions. Finally, though people’s within-city travel distance did not have a large impact on the number of cases, it is important for shaping spatial patterns, which can have implications for control activities and for local herd immunity.

## Acknowledgements

We would like to acknowledge Dr. Jason R. Andrews for the valuable input in this work. We would also acknowledge the fieldwork teams (in Kisumu: Joel Mbakaya, Samwel Ndire and Charles Adipo; in Ukunda: Said Lipi Malumbo, Paul S. Mutuku, Charles M. Ng’ang’a) who collected the data that made this work possible.

## Data availability statement

All the data, codes, and resources to recreate the simulations for the original model and the modification reported in this work are publicly available at https://github.com/vhpenagarcia/ABM_dengue and have been archived within the Zenodo repository (https://doi.org/10.5281/zenodo.14036270).

