## Supporting information for "Mobility and non-household environments: understanding dengue transmission patterns in urban contexts"

### Supporting Methods

#### Mosquito movement parameterization

We consider two variables to estimate a baseline migration probability for the number of mosquitoes moving from given locations: availability of both breeding and blood-feeding resources.

The model includes a local density-dependent function that allows for the mosquito population to grow in a location-specific way, allowing it to represent dynamics previously described for a fragmented environment.[1] By using this function, the number of mosquitoes in each location grows according to availability of water containers, the density of immature mosquitoes in those containers, and temperature (peaking at 29°C), as follows:

$$f(D) = \frac{1}{1 + e^{0.09D - 0.55}} d(T) \tag{1}$$

Where

$$d(T) = -0.166 + 0.08T - 0.0014T^2 \tag{2}$$

In the equation, $D$ is the larval density expressed as the ratio of the number of larvae to liters of water available for breeding in the structure, and $d(T)$ is the term describing the temperature-dependence of population growth. The equation allows the sub-population to grow when larval density is low by assuming they occupy all the water containers inside or around the structure. As the mosquito sub-population grows, resource availability in the water declines, slowing population growth. By considering $Nm_t$ as the mosquito subpopulation size at time $t$, when the ratio $Nm_t/Nm_{t+1}$ approaches 1, breeding site resources are reaching their carrying capacity and hence, the probability of a mosquito migrating increases. Because there is a strong stochastic component in estimating $Nm_{t+1}$, by running 20 simulations for every location every day, we estimated a parameter $MP$ as the median of ratios $Nm_t/Nm_{t+1}$.

We also consider the probability that each female mosquito bites a human within a given location. The rate of biting is temperature dependent ($a$), according to previously published work.[2] We coupled it into a binomial distribution to define the number of biting mosquitoes as follows:

$$Nb \sim Bin(Nm_t, a) \tag{3}$$

Where $Nb$ is the number of females needing to bite on day $t$ and $Nm_t$ is the size of sub-population mosquitoes that day. While most models assume that all females bite when they need to, here we consider the possibility that mosquitoes cannot find blood meal hosts in a given place and time as dependent on the human occupancy of a given location. We assume that there is a probability for $Nb$ mosquitoes to have a successful biting human-mosquito encounter ($P(bite)$) based on the size of $Nb$ and the amount of time that humans spend in the location, as follows:

$$P(bite) = 1 - e^{-\frac{h}{24}Nh_t} \tag{4}$$

Where $Nh_t$ is the number of people attending a given location on day t for a structure-specific number of hours. Finally, the actual number females that fed ($NF$) in each day, is estimated by

$$NF \sim Bin\big(Nb, P(bite)\big) \tag{5}$$

In this sense, when the number of people on a given day is low and the number of hours they spend in the location are few, the probability of a female mosquito successfully biting is low. So, as the ratio $NF/Nb$ (later called $MF$) approaches 0, the probability of mosquitoes migrating increases. Again, for every location every day, 20 simulations were run to estimate a distribution of $MF$.

Finally, we estimated the migration probability as the product $Mp \cdot (1\text{-}MF)$, which depends on both larval density conditions via $Mp$ and adult biting conditions via $MF$. The resulting probability was used in a binomial distribution to determine the number of migrating mosquitoes each day. To evaluate the effect of different mosquito movement regimes on the burden of dengue, we considered migration rates of 100% (as unmodified product $Mp \cdot (1\text{-}MF)$), 50%, and 10%. The total number of cases was quantified after 200 simulations.

Once a mosquito migrates, a new location is assigned by considering a dispersal kernel [3]. Following previous work,[4] we used a lognormal function with the form

$$PD = \frac{1}{(2\pi)^{\frac{3}{2}} b d^2} e^{-\frac{\log\left({}^{d}/_{a}\right)^2}{2b^2}} \tag{6}$$

Where $d$ is distance and both $a$ and $b$ are parameters to be estimated. We fitted a function by assuming a mean dispersal distance of 105.69 meters, as estimated for *Aedes aegypti* by.[5]

#### Use of temperature-dependent function into the model

Biological and transmission-related traits of mosquitoes are strongly dependent on temperature. The mathematical formulations and their implementation within the model are described in the following table as reported previously [6]:

**Table S1: Implementation of mosquito-related functions to model population dynamics traits.** Functions were extracted from previous works originally fitted by Mordecai [2] and later modified and used by Huber [7] and Caldwell [8]. Functions can be either Brière [$cT(T-T_{min})(T_{max}-T)^{1/2}$] or quadratic [$c(T-T_{max})(T-T_{min})$].

| Trait | Estimation function | | | | Use |
|---|---|---|---|---|---|
| | function | c | Tmin | Tmax | |
| Human biting rate (*a*) | Brière | 2.02e$^{-04}$ | 13.35 | 40.08 | $N_B \sim Bin(Nm, a)$* |
| Mortality rate (*μ*) | Quadratic | -1.48e$^{-01}$ | 9.16 | 37.73 | $N_D \sim Bin(Nm, \mu)$+ |
| Probability of infection of a mosquito (*c*) | Brière | 4.91e$^{-04}$ | 12.22 | 37.46 | $vc = b \cdot c$† <br> $E \sim Bin(N_{bit}, vc)$‡ |
| Probability for a mosquito to become infectious (*b*) | Brière | 8.49e$^{-04}$ | 17.05 | 35.83 | |
| Parasite development rate (*PDR*) | Brière | 6.65e$^{-05}$ | 10.68 | 45.90 | $inf \sim Bin(E, PDR)$¥ |
| Eggs per female per day (*EFD*) | Brière | 8.56e$^{-03}$ | 14.58 | 34.61 | $Eggs \sim Poisson(EFD)$ |
| Mosquito development rate (*MDR*) | Brière | 7.86e$^{-05}$ | 11.36 | 39.17 | $r = MDR \cdot pEA \cdot f(D)$ <br> $Em \sim Bin(N_L, r)$§ |
| Egg-to-adult survival probability (*pEA*) | Quadratic | -5.99e$^{-03}$ | 13.56 | 38.29 | |

* $N_B$, the number of mosquitoes at a given location biting that specific day.

+ $N_D$, the number of adult mosquito deaths happening in a given day at a specific location.

† Parameters $b$ and $c$ are the components of vector competence ($vc$).

‡ $E$, number of exposed mosquitoes, $N_{bit}$ refers to those mosquitoes that bit an infected individual, so they are moved to exposed infection status with a probability $vc$.

¥ *inf* refers to the moment when a mosquito is moved to infectious stage which is determined by a rate $PDR$

§ $Em$, number of emerged mosquitoes, $N_L$ is the number of larvae at a given location.

### Supporting Results

#### Movement of mosquitoes is costly and reduces human dengue burden

When we considered different regimes of mosquito movement, the model yielded a lower number of cases when mosquito migration was higher. The median number of cases was 306 (IQR: 222 – 419) at 100% of mosquito movement and increased to 1,090 (IQR: 533 – 1,714) and 4,670 (IQR: 4,005 – 5,229) when number of mosquitoes was 50% and 10%, respectively (Table S4). Still the proportion of infections happening in HH compared to NH remained lower regardless of mosquito movement (Fig S8). To better understand the mechanism by which mosquito movement was reducing transmission, we studied the size of mosquito populations for each mosquito movement regime, which resulted in smaller population sizes at higher movement rate (Fig S9), indicating the importance of mosquito mortality during migration as a limit on transmission.

### Supporting Discussion

In this model we also included estimates of mosquito movement, where we found that high levels of mosquito migration induce high mosquito mortality, thereby reducing mosquito abundance and transmission (Fig S8 and S9). These results suggest that mosquito migration is carried out at a high cost for the individual and population, which supports the empirical observation that mosquitoes tend to stay in or close to the same location where they are breeding. Though our model has different parameterization and purpose from that previously reported by Reiner and colleagues, they also included both movement of people and mosquitoes and found that the former is the real force shaping dengue transmission as opposed to movement of mosquitoes, which diffuses it [9]. Interestingly, they found that their model output matches real observations when mosquito movement is decreased and, under certain scenarios, when mosquito movement equals zero [9].

In our model, a migrant mosquito travels to another location, irrespective of the suitability of that location (i.e., presence of water containers or established mosquito population), with a distance-based probability. If the new location is suitable for breeding, a new mosquito subpopulation is established, otherwise the migrant mosquito will die before reproducing. A migrant mosquito may be more attracted to travel to a location with water containers, as suggested by other studies [10], which is not included in the model. However, it is not known how likely it is for a migrating mosquito to travel longer distances in search of containers during migration if the closest location does not have any. This type of process has never been studied before. Another variable not considered is the number of buildings, roads, and other urban features that might impose some limits to mosquito dispersal like those described previously, where all *Ae. aegypti* individuals were recaptured in the same block where they were released, unable to cross roads [11].

Though there are several mark-release-recapture studies recording long travel distances by *Ae. aegypti* [5], most of these studies are artificially releasing mosquitoes where they are forced to travel looking for a place to settle. While these studies are useful to evaluate the capacity of mosquitoes to fly in these conditions, they are not indicative of the propensity for already settled mosquitoes to travel the distances recorded in these studies.

According to results of this model, intra-urban dispersion of dengue is not explained by mosquito migration. A plausible explanation is that dispersion is actively driven by human mobility. This would underscore the importance of human movement by suggesting that it is mainly responsible for the burden and dispersal of dengue through communities. Though the role of human mobility on intra-urban mosquito dispersion is hard to measure, there are several studies explaining the importance of human movement in dengue transmission at different spatial levels [9, 12-17].

### Supporting Figures

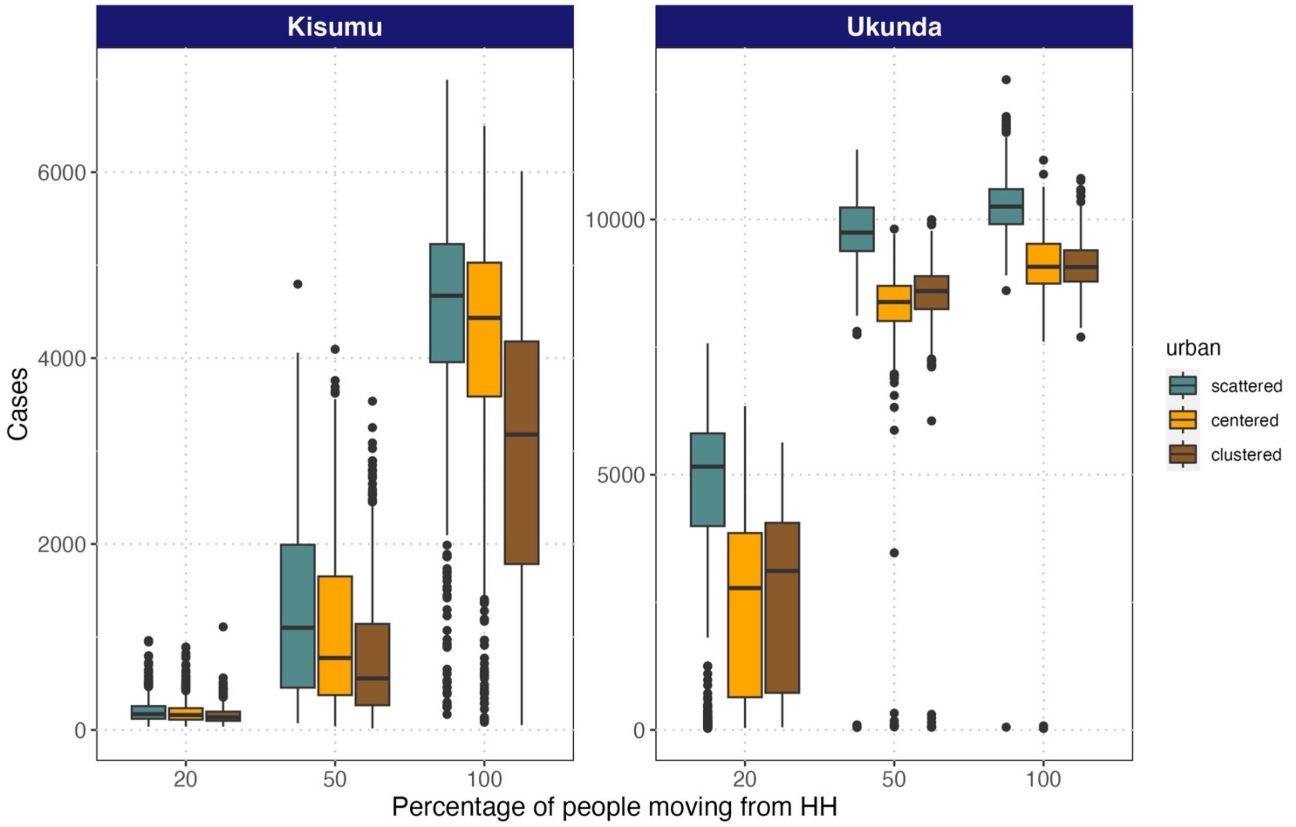


**Fig S1: Decreasing the number of people moving to NH from HH decreases the number of infections.** A higher burden of dengue at scattered distribution of NH is also evident for both Kenyan cities of Kisumu and Ukunda. The horizontal line indicates the median of 200 runs, while box represents interquartile range (IQR). Whiskers show the range of 1.5·IQR extending beyond the box. Dots are data points outside the whole range.

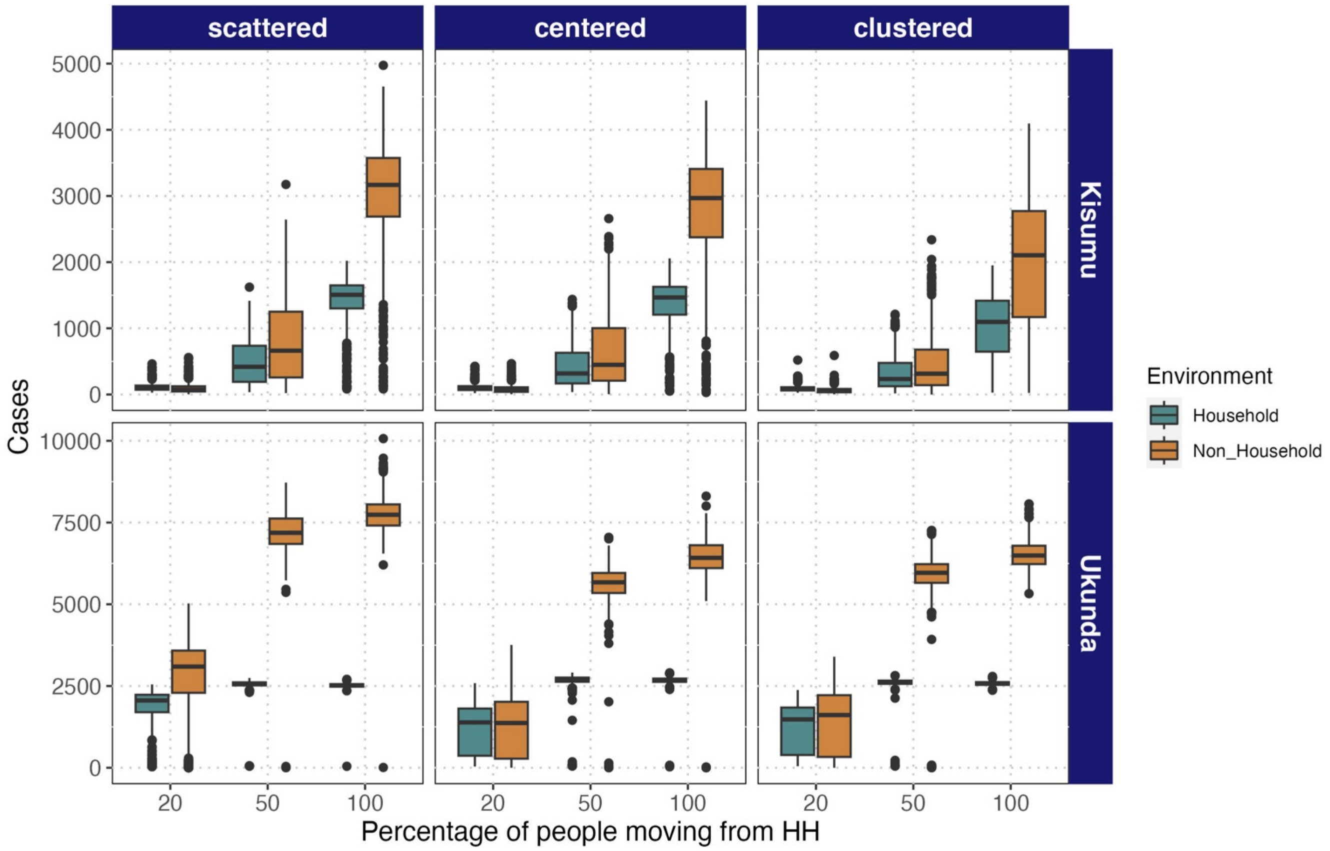


**Fig S2: Decreasing the number of people moving from HH to NH environments decreases the burden of dengue, which is mediated by infections produced in NH.** The boxes show the IQR of cases of 200 runs for each simulation, median is represented by horizontal line, whiskers represent the range beyond the box of 1.5·IQR, and the dots are data points outside such range.

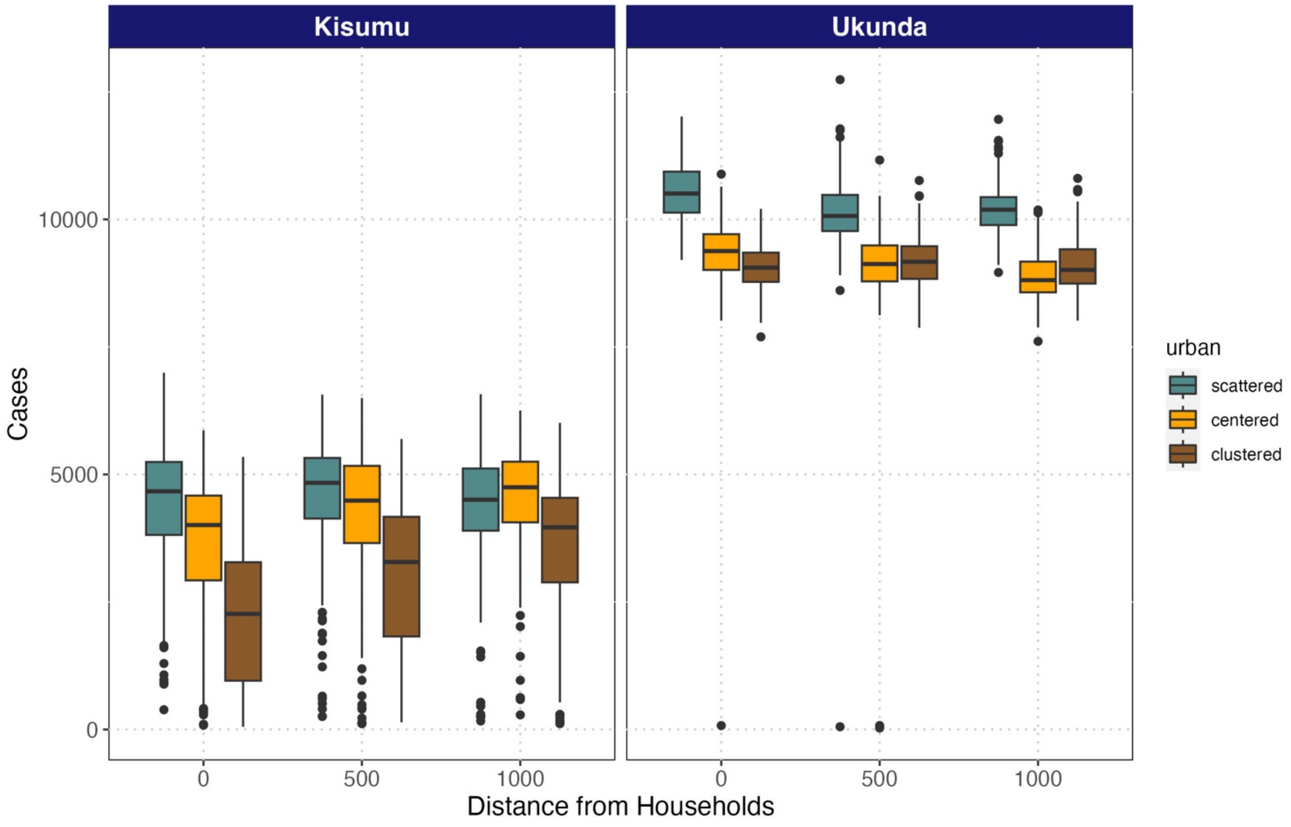


**Fig S3: Burden of dengue in Kisumu and Ukunda is city-specific and not greatly modified by distance that people travel from HH to NH.** A higher burden is achieved when NH are randomly spatially distributed (scattered). Three levels of distance were assessed (the closest NH from HH [categorized as 0], at least 500 meters, and at least 1000 meters).

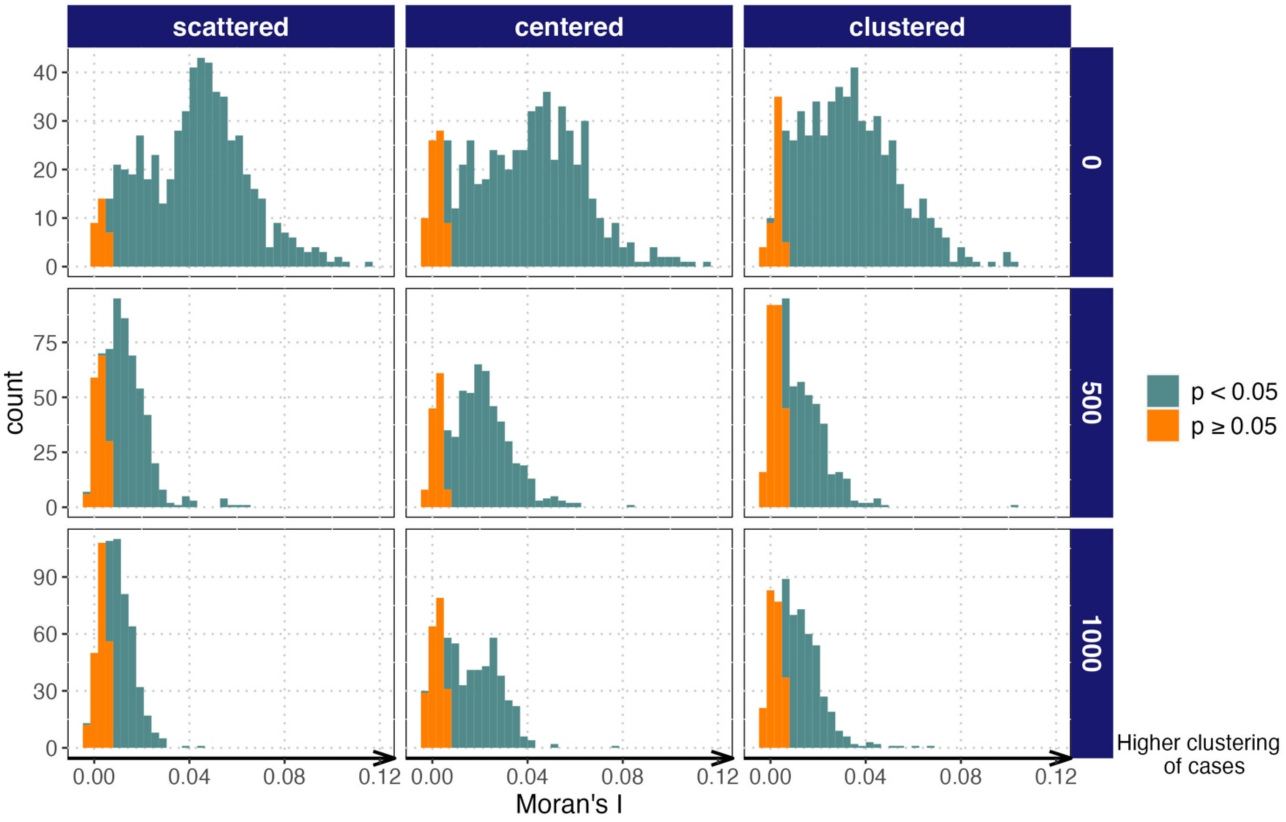


**Fig S4: Moran's $I$ value increases as distance travelled by people between HH and NH decreases across different urban configurations in Kisumu.** Distribution of values are shown across three different urban conformations and different distance regimes (distance traveled by people from HH to NH: the closest distance [categorized as zero], and at least 500 and 1000 meters). Significance with $\alpha$ = 0.05 is shown according to color. The level of clustering of dengue cases increases with Moran's $I$ value.

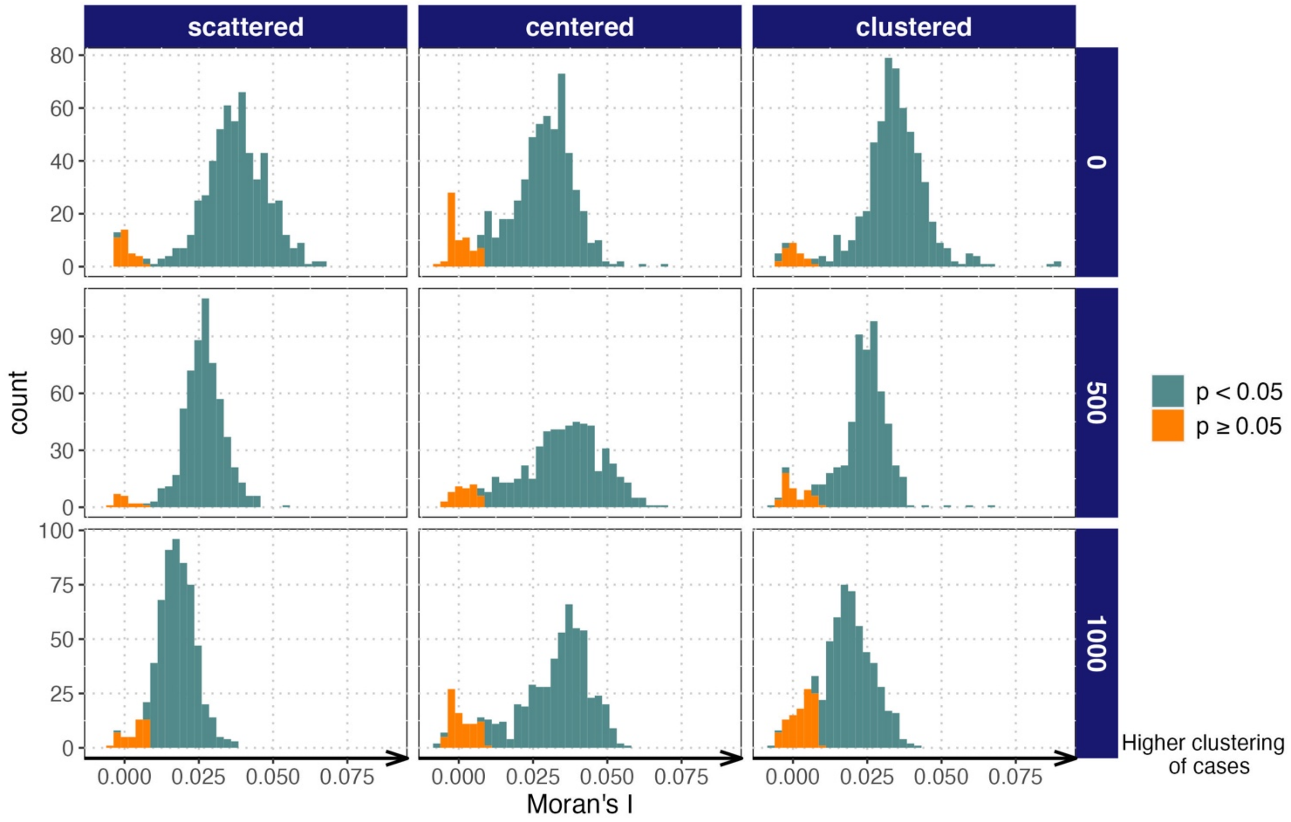


**Fig S5: Moran's $I$ value increases as distance travelled by people between HH and NH decreases across different urban configurations in Ukunda.** Distribution of values are shown across three different urban conformations and different distance regimes (distance traveled by people from HH to NH: the closest distance [categorized as zero], and at least 500 and 1000 meters). Significance with $\alpha$ = 0.05 is shown according to color. The level of clustering of dengue cases increases with Moran's $I$ value.

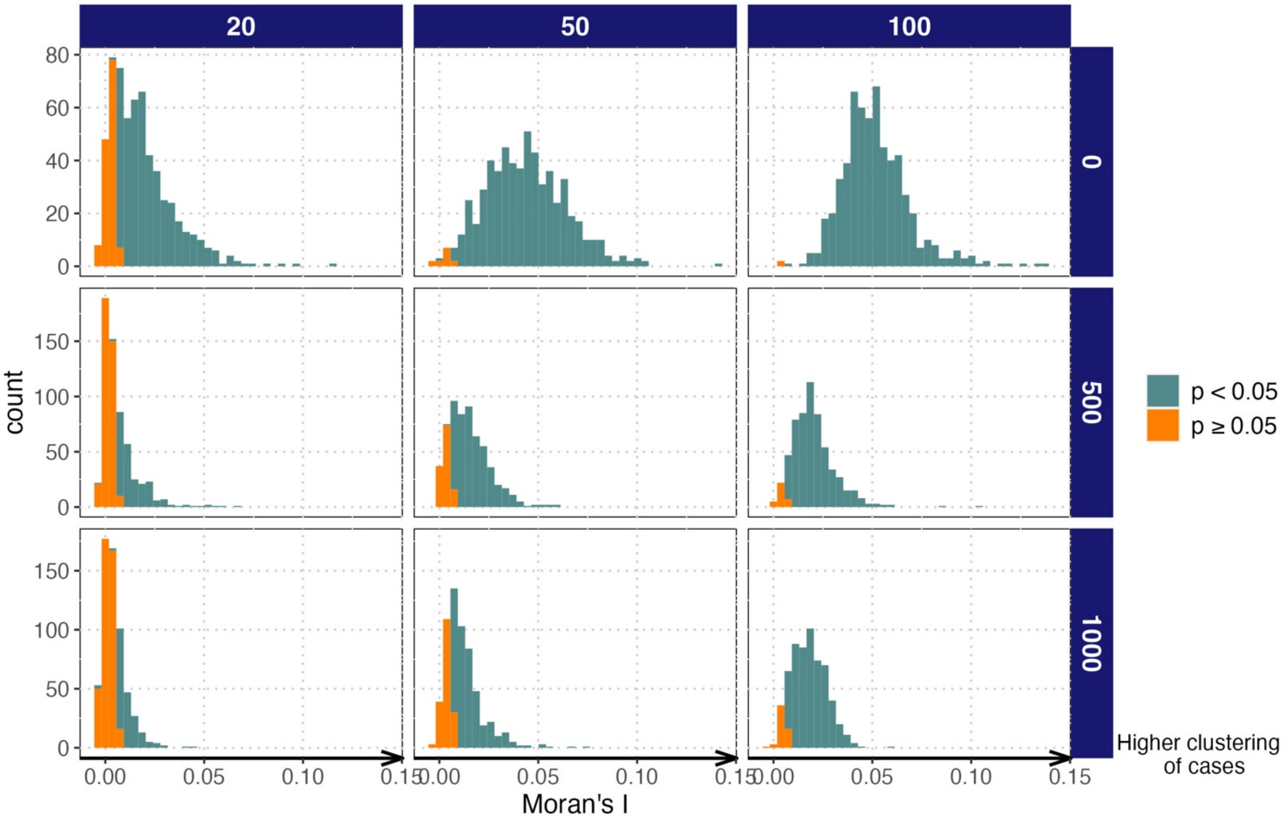


**Fig S6: Moran's I value increases as number of people travelling to NH increase and distance travelled by people between HH and NH decreases in Kisumu.** Distribution of values are shown across three different percentages of people travelling and different distance regimes (distance traveled by people from HH to NH: the closest distance [categorized as zero], and at least 500 and 1000 meters). Significance with $\alpha = 0.05$ is shown according to color. The level of clustering of dengue cases increases with Moran's $I$ value.

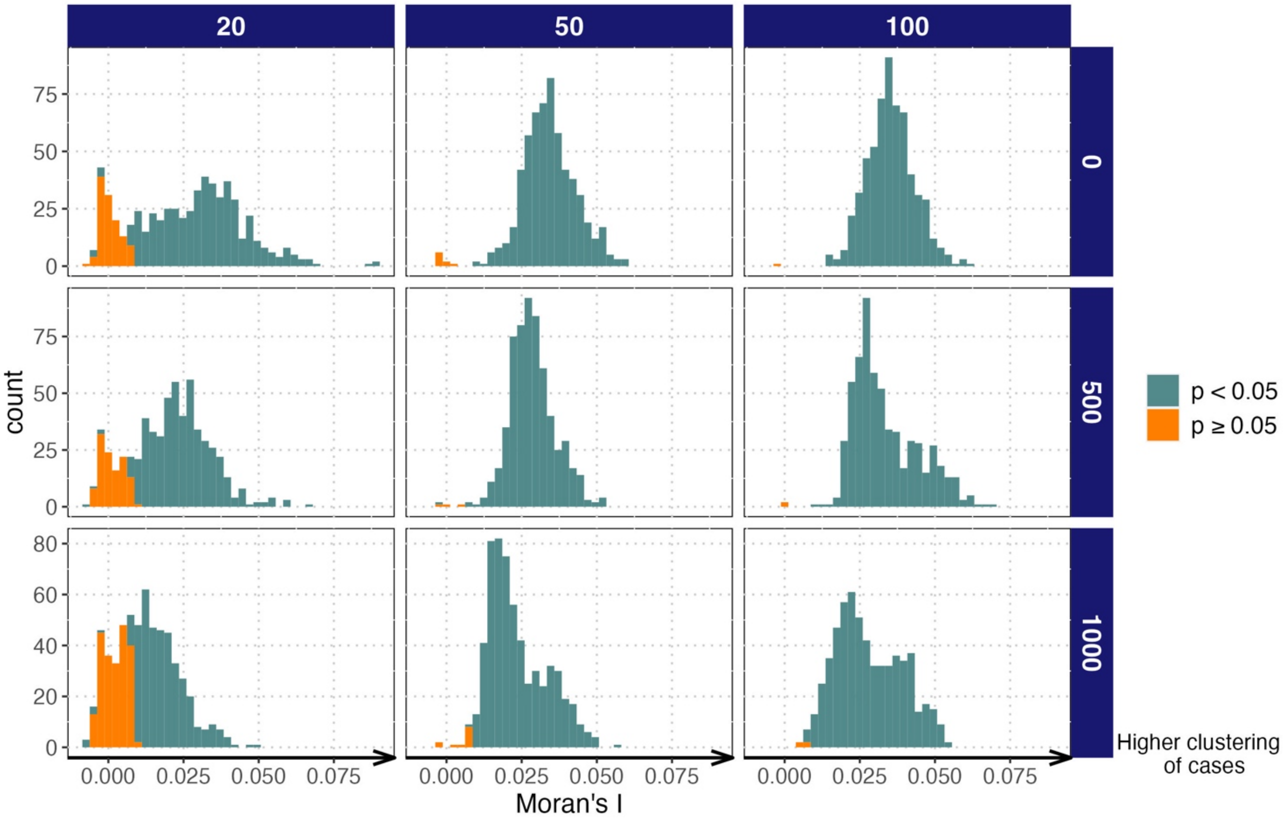


**Fig S7: Moran's I value increases as number of people travelling to NH increase and distance travelled by people between HH and NH decreases in Ukunda.** Distribution of values are shown across three different percentages of people travelling and different distance regimes (distance traveled by people from HH to NH: the closest distance [categorized as zero], and at least 500 and 1000 meters). Significance with $\alpha$ = 0.05 is shown according to color. The level of clustering of dengue cases increases with Moran's $I$ value.

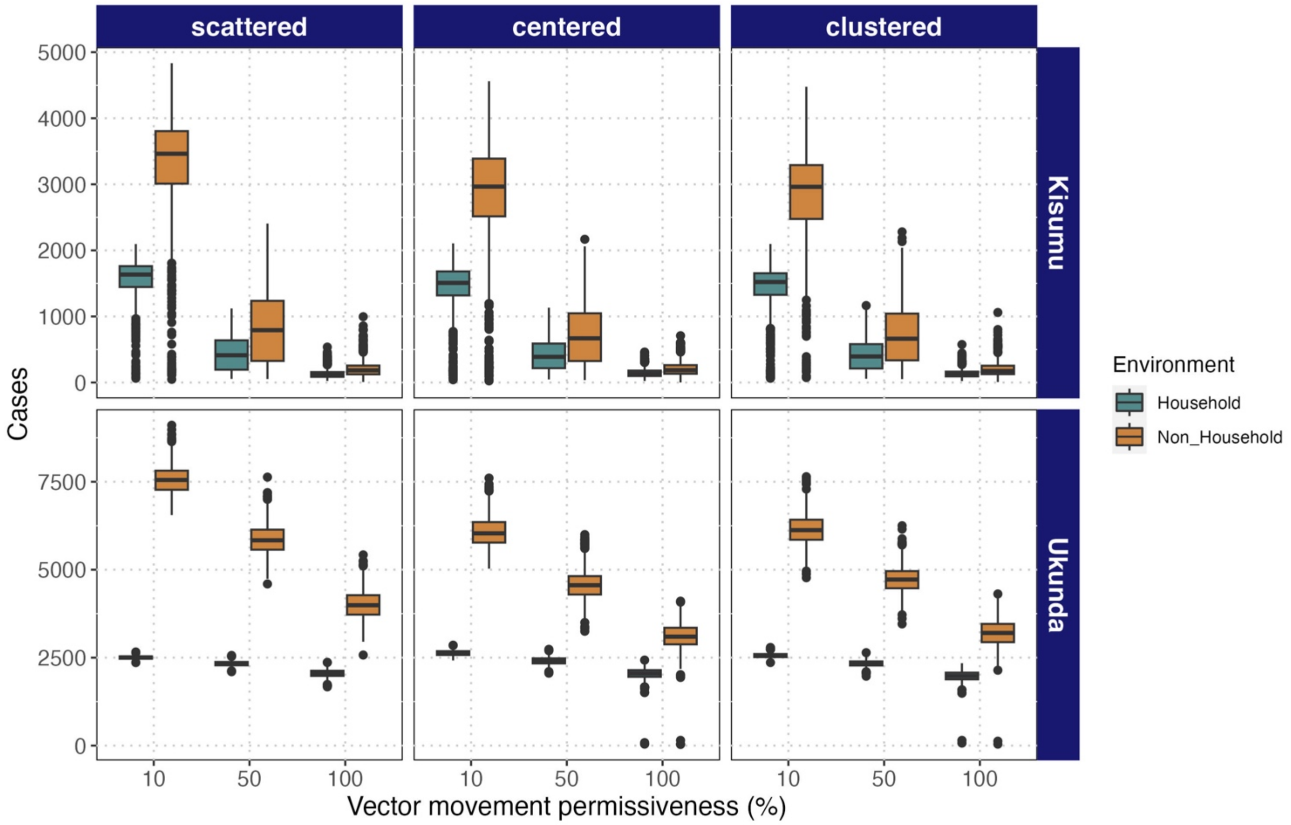


**Fig S8: The number of infections happening in both environments is affected by the level of mosquito movement.** Difference on the number of infections happening in HH and NH and in turn the burden of dengue decreases as mosquito movement decreases as well. Boxplots are showing the distribution of the number of cases for three levels of mosquito movement (x axis) and three urban conformations (top panels) for two Kenyan cities (Kisumu and Ukunda).

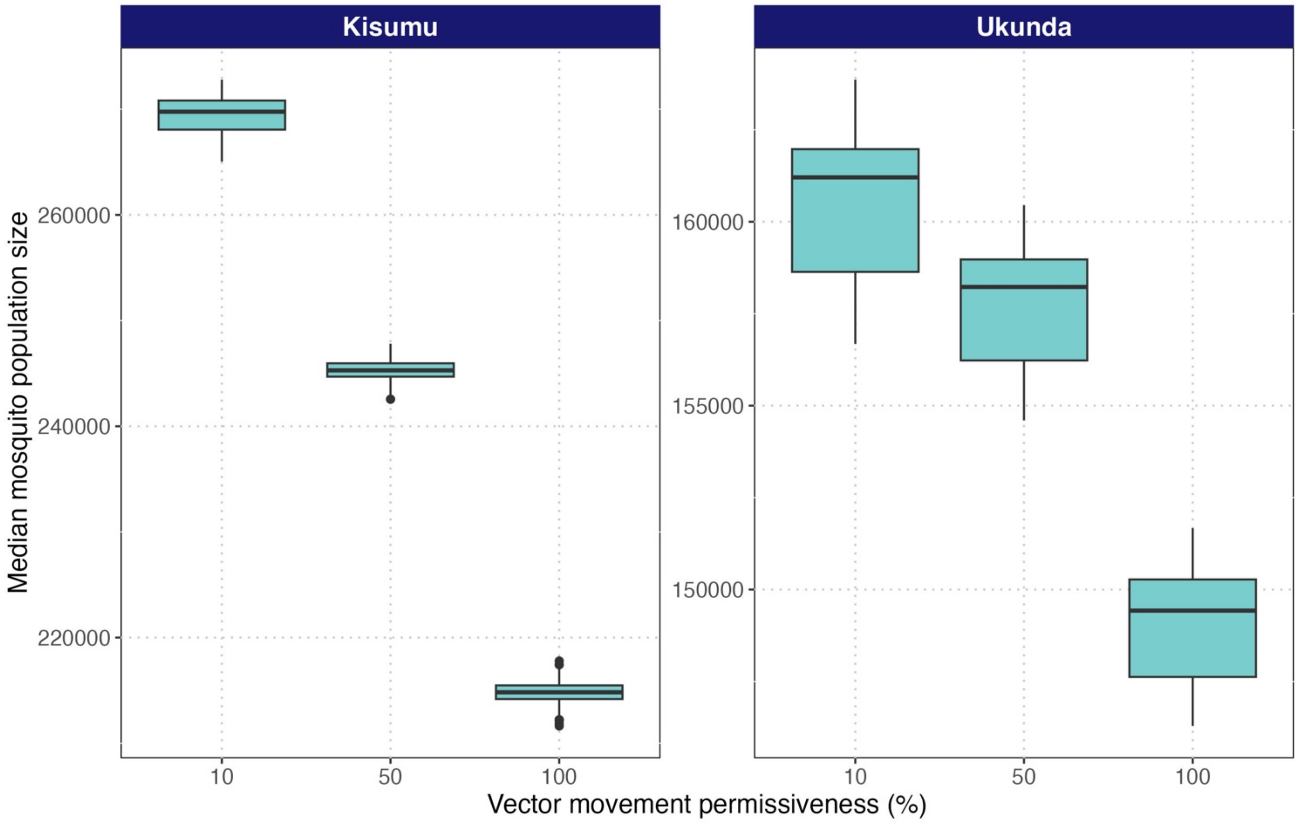


**Fig S9: Mosquito population sizes across three levels of vector movement evidenced the decrease of mosquito populations when vector movement is increasing.** Boxplots show the distribution of the median size of population size across the two modelled years for 200 runs for two Kenyan cities (Kisumu and Ukunda).

#### Supporting tables

**Table S2: Descriptive outcomes for 200 runs at different level of human movement from HH to NH in Kenyan cities of Kisumu and Ukunda.** Three levels of human movement were assessed: 100% (base level of movement), 50%, and 20%. Descriptive values for 200 runs include the median of the total number of infections, interquartile range (IQR), and the proportion of infections that occur in any of the five different types of NH environments.

| City | Human movement's level | Median | IQR | Proportion of infections in NH |
|---|---|---|---|---|
| Kisumu | 20 | 154 | 108 – 232 | 0.423 |
| | 50 | 764 | 349 – 1626 | 0.588 |
| | 100 | 4228 | 3025 – 4921 | 0.670 |
| Ukunda | 20 | 3544 | 1416 – 4716 | 0.531 |
| | 50 | 8770 | 8305 – 9428 | 0.699 |
| | 100 | 9416 | 8923 – 10028 | 0.724 |

**Table S3: Descriptive outcomes for 200 runs at different urban conformations of non-household environments for Kenyan cities of Kisumu and Ukunda.** Three urban conformations were assessed: Scattered (when non-household (NH) environments is randomly distributed in space), centered (when most of NH are grouped in the center), and clustered (when NH are grouped in three clusters). Descriptive values for 200 runs include the median of the total number of infections, interquartile range (IQR), and the proportion of infections that occur in any of the five different types of NH environments.

| City | Urban conformation | Median | IQR | Proportion of infections in NH |
|---|---|---|---|---|
| Kisumu | Scattered | 4672 | 3956 – 5227 | 0.678 |
| | Centered | 4432 | 3587 – 5027 | 0.669 |
| | Clustered | 3178 | 1785 – 4179 | 0.657 |
| Ukunda | Scattered | 10254 | 9909 – 10596 | 0.754 |
| | Centered | 9074 | 8744 – 9524 | 0.706 |
| | Clustered | 9067 | 8784 – 9398 | 0.716 |

**Table S4: Distance traveled by people from HH to NH have little effect on total burden of dengue.** Three levels of intra-urban distance from HH to NH were evaluated: the closest NH location from HH (categorized as zero), at least 500 meters away, and at least 1000 meters away. The table shows the descriptive outcomes for4 200 runs which include the median of the number of cases, the interquartile range (IQR), and the proportion of infections happening in any of the five different types of NH environments.

| **City** | **Distance traveled from HH to NH (meters)** | **Median** | **IQR** | **Proportion of infections in NH** |
|---|---|---|---|---|
| **Kisumu** | 0 (closest) | 3,742 | 2,320 – 4,671 | 0.662 |
| | 500 | 4,284 | 3,170 – 5,028 | 0.674 |
| | 1000 | 4,432 | 3,642 – 4,992 | 0.669 |
| **Ukunda** | 0 (closest) | 9,534 | 9,024 – 10,255 | 0.726 |
| | 500 | 9,408 | 8,958 – 9,942 | 0.724 |
| | 1000 | 9,269 | 8,785 – 9,965 | 0.721 |

**Table S5: Descriptive outcomes for 200 runs at different levels of vector movement for Kenyan cities of Kisumu and Ukunda**. Three levels of vector movement were assessed: 100%, 50%, and 10%. Descriptive values for 200 runs include the median of the total number of infections, interquartile range (IQR), and the proportion of infections that occur in any of the five different types of NH environments.

| **City** | **Vector movement's level (%)** | **Median** | **IQR** | **Proportion of infections in NH** |
|---|---|---|---|---|
| Kisumu | 10 | 4670 | 4005 – 5229 | 0.667 |
| | 50 | 1090 | 533 – 1714 | 0.663 |
| | 100 | 306 | 222 – 419 | 0.592 |
| Ukunda | 10 | 9007 | 8536 – 9795 | 0.714 |
| | 50 | 7306 | 6859 – 7961 | 0.676 |
| | 100 | 5428 | 4991 – 5911 | 0.625 |
